# Retention and loss to follow-up among patients with hypertension in primary care: a multi-practice cohort study

**DOI:** 10.64898/2026.05.10.26352856

**Authors:** Jiancheng Ye, Alan Song

## Abstract

Effective hypertension management depends on sustained engagement with primary care, and there is a need to understand the magnitude and determinants of follow-up loss in real-world primary care. We analyzed electronic health record (EHR) data from 26,541 patients with hypertension across primary care practices participating in the EvidenceNOW quality-improvement initiative. We characterized retention in care, longitudinal blood pressure (BP) control, and predictors of loss to follow-up using descriptive statistics, cumulative retention curves, and multivariable Cox proportional-hazards regression. At baseline, mean systolic and diastolic BP were 140.0 ± 20.6 and 84.7 ± 13.0 mmHg, respectively; only 10.7% (95% CI 10.4–11.1) of patients had controlled BP and 18.1% never returned for any follow-up visit. Among the 21,729 patients who had ≥1 follow-up encounter, retention declined steeply over time—from 59.9% at 6 months to 16.3% at 36 months. Patients identifying as Black/African American (adjusted hazard ratio [aHR] 1.44; 95% CI 1.33–1.56), Hispanic/Latino (aHR 1.43; 1.35–1.52), or Other race/ethnicity (aHR 1.50; 1.41–1.59) had significantly higher hazards of being lost to follow-up than White patients, whereas older age, female sex, comorbid diabetes, heart failure, chronic kidney disease, stroke, and baseline BP control were each independently protective. Among patients retained for at least 12 months, BP control rose to 63.7% and remained near 64–66% through 36 months. These findings reveal a substantial and inequitable longitudinal care-engagement gap that is likely a principal driver of suboptimal hypertension control in the United States and identify actionable demographic and clinical targets for primary-care retention interventions.

## INTRODUCTION

Hypertension is the leading modifiable cause of cardiovascular morbidity and mortality worldwide and remains the single largest contributor to all-cause death in the United States.[1] Despite decades of investment in evidence-based guidelines, low-cost generic therapies, and population-level screening campaigns, fewer than one in three U.S. adults with hypertension achieves blood pressure (BP) at target, and recent national surveillance data suggest control rates have plateaued or declined since 2017.[2] Closing this gap is widely regarded as one of the highest-yield interventions available to public health.[3, 4]

Hypertension is fundamentally a chronic condition whose management depends on the sustained therapeutic relationship between a patient and a primary-care team: titration of pharmacotherapy, reinforcement of lifestyle counseling, and surveillance for end-organ injury all require repeated longitudinal contact.[5, 6] A growing body of work has therefore shifted attention from the question of whether the right medications are prescribed at a single encounter to whether patients remain engaged in care long enough for those medications—and the broader plan—to take effect.[7, 8] Yet the structure of fee-for-service primary care, social determinants of health such as transportation and language access,[9] and competing demands of multimorbidity all conspire to make sustained engagement difficult.[10] Studies of large integrated health systems have hinted at substantial loss to follow-up among hypertensive patients,[11] but data from community primary-care practices that serve diverse, lower-income populations in the United States are sparse, and few have linked engagement patterns to longitudinal BP outcomes within the same cohort.[12]

EvidenceNOW is a federally funded primary-care quality-improvement program that supports small-to-medium primary-care practices in delivering evidence-based cardiovascular preventive care.[13] The program’s standardized electronic health record (EHR) data offer a unique opportunity to characterize how patients with hypertension actually move through real-world primary care over time, and to examine which patient and clinical characteristics predict disengagement. Here we describe a multi-practice cohort of 26,541 patients with hypertension, quantify retention in care across 36 months of follow-up, characterize longitudinal BP control among those retained, and identify demographic and clinical predictors of loss to follow-up using multivariable survival analysis. We report the findings to inform the design of equitable, retention-focused interventions that can complement existing treatment-intensification efforts and translate prescribing improvements into population-level BP control.[14]

## METHODS

### Data source

This study used deidentified EHR data from primary-care practices participating in the EvidenceNOW quality-improvement program, which supports small-to-medium primary-care practices across multiple U.S. regions in delivering evidence-based cardiovascular preventive care.[13] Standardized data extracts included diagnoses (ICD codes and SNOMED CT terms), enabling identification of hypertension and key comorbidities including type 2 diabetes mellitus, coronary artery disease, prior myocardial infarction, heart failure, chronic kidney disease, and stroke; demographics (age at diagnosis, sex, self-reported race and ethnicity, marital status, and preferred language for healthcare communication); vital signs at each encounter (systolic and diastolic BP, heart rate, body mass index [BMI], and body weight); and medication records (medication name, standardized codes, dosing frequency, and start/end dates). The study used only deidentified secondary data and was determined to be non-human-subjects research by the relevant institutional review board.

### Study population and patient selection

Eligible patients were adults aged ≥18 years with a documented hypertension diagnosis and at least one primary-care encounter in the EvidenceNOW database. We extracted longitudinal BP histories for all eligible patients and computed derived variables including mean, maximum, and minimum systolic and diastolic BP across visits; total visit count; duration between consecutive visits; days since most recent visit; and visit-interval variability. These longitudinal features were merged with demographic and comorbidity data to create the analytic dataset. Patients with at least two clinical visits were used for analyses requiring longitudinal vital-sign trajectories; the full cohort (including patients with only an index visit) was used for retention analyses to avoid conditioning on the outcome.

### Measurements definitions

Baseline was defined as the index hypertension encounter. Follow-up was defined as any subsequent primary-care encounter. Retention at t months was defined as the presence of at least one encounter occurring after time t from the index visit, calculated only among patients with sufficient potential observation time at that landmark. BP control was defined as systolic BP <140 mmHg and diastolic BP <90 mmHg, consistent with widely used quality-measure thresholds.[5] Treatment was defined as an active prescription for at least one antihypertensive medication on the encounter date. Loss to follow-up was the time-to-event outcome for the survival model and was defined as the date of the last observed encounter for patients without subsequent contact through the end of the observation window.

### Statistical analysis

Continuous variables are presented as mean ± standard deviation or median (interquartile range) and compared between groups using Welch’s t-test or Wilcoxon rank-sum test as appropriate. Categorical variables are presented as counts and percentages and compared using chi-square or Fisher’s exact tests. The 95% percent confidence intervals (CI) for proportions were computed using the Clopper–Pearson exact method. Cumulative retention curves were estimated using a Kaplan–Meier-style approach: patients contributed time from their index visit until their last observed encounter, and the time-to-event outcome was the time at which a patient was last seen in primary care, with patients censored at the end of the observation window if they had a recent encounter consistent with ongoing engagement.

Multivariable Cox proportional-hazards regression was used to identify independent predictors of loss to follow-up. Candidate covariates were prespecified and included age (per 1-SD increase, continuous), sex, race/ethnicity (referent: White), BMI category (referent: normal weight 18.5–24.9 kg/m²), comorbid diabetes, heart failure, chronic kidney disease, stroke, baseline BP control status, and baseline antihypertensive treatment status. The proportional-hazards assumption was assessed using scaled Schoenfeld residuals; no material violations were detected. Adjusted hazard ratios are reported with 95% confidence intervals. All p-values are two-sided, with statistical significance defined as p < 0.05. Analyses were performed using Python (version 3.11) with the pandas, statsmodels, and lifelines packages, and R (version 4.3) for survival visualization.

### Study Approval

This study utilized the dataset from a quality improvement program (EvidenceNOW). All the data were de-identified and the study was approved by Northwestern University’s Institutional Review Board.

## RESULTS

### Cohort characteristics

We identified 26,541 patients with a documented diagnosis of hypertension and at least one primary-care encounter in EvidenceNOW practices during the study period (**Table 1**). The mean age was 54.3 ± 14.0 years; 56.2% were female; the cohort was racially and ethnically diverse, with Hispanic/Latino patients comprising 35.6%, White patients 7.5%, Black/African American patients 3.6%, Asian patients 2.7%, and 50.5% reporting another or unknown race/ethnicity, reflecting both the population served by participating practices and known limitations of EHR race/ethnicity capture. English (50.6%) and Spanish (25.0%) were the most commonly reported preferred languages. Comorbid diabetes was present in 47.1% of patients, chronic kidney disease in 3.7%, heart failure in 4.0%, prior stroke in 4.9%, and prior myocardial infarction in 3.5%. Lisinopril (24.9%), aspirin (23.0%), metformin (17.0%), atorvastatin (15.6%) and hydrochlorothiazide (15.0%) were the most frequently prescribed agents.

**Table 1.** Baseline characteristics of the hypertension cohort, overall and stratified by follow-up status.

| Characteristic | Overall cohort (N = 26,541) | No follow-up visit (N = 4,811) | ≥1 follow-up visit (N = 21,729) | p-value |
| --- | --- | --- | --- | --- |
| Age, mean ± SD, years | 54.3 ± 14.0 | 54.7 ± 14.3 | 54.2 ± 14.0 | 0.03 |
| **Age group, n (%)** |  |  |  |  |
| 18–39 years | 3,996 (15.1) | 750 (15.6) | 3,246 (14.9) | 0.265 |
| 40–64 years | 17,166 (64.7) | 3,038 (63.1) | 14,128 (65.0) | 0.014 |
| ≥65 years | 5,276 (19.9) | 1,016 (21.1) | 4,260 (19.6) | 0.019 |
| **Sex, n (%)** |  |  |  |  |
| Female | 14,912 (56.2) | 2,640 (54.9) | 12,272 (56.5) | 0.043 |
| Male | 11,629 (43.8) | 2,172 (45.1) | 9,457 (43.5) | — |
| BMI, mean ± SD, kg/m<sup>2</sup> (n with data) | 30.9 ± 7.3 (5,489) | 30.3 ± 7.1 (666) | 31.0 ± 7.3 (4,823) | 0.02 |
| **Race/ethnicity, n (%)** |  |  |  |  |
| White | 2,000 (7.5) | 223 (4.6) | 1,777 (8.2) | <0.001 |
| Black/African American | 943 (3.6) | 159 (3.3) | 784 (3.6) | 0.324 |
| Hispanic/Latino | 9,460 (35.6) | 1,454 (30.2) | 8,006 (36.8) | <0.001 |
| Asian | 722 (2.7) | 61 (1.3) | 661 (3.0) | <0.001 |
| Other/Unknown | 13,416 (50.5) | 2,915 (60.6) | 10,501 (48.3) | <0.001 |
| **Preferred language, n (%)** |  |  |  |  |
| English | 13,430 (50.6) | 2,814 (58.5) | 10,616 (48.9) | <0.001 |
| Spanish | 6,636 (25.0) | 1,100 (22.9) | 5,536 (25.5) | <0.001 |
| Other | 647 (2.4) | 130 (2.7) | 517 (2.4) | 0.208 |
| Missing/Unknown | 5,828 (22.0) | 768 (16.0) | 5,060 (23.3) | <0.001 |
| **Comorbidities, n (%)** |  |  |  |  |
| Diabetes | 12,506 (47.1) | 1,568 (32.6) | 10,938 (50.3) | <0.001 |
| Heart failure | 1,054 (4.0) | 160 (3.3) | 894 (4.1) | 0.013 |
| Stroke | 1,299 (4.9) | 115 (2.4) | 1,184 (5.4) | <0.001 |
| Prior myocardial infarction | 937 (3.5) | 121 (2.5) | 816 (3.8) | <0.001 |
| Chronic kidney disease | 994 (3.7) | 117 (2.4) | 877 (4.0) | <0.001 |
| **Antihypertensive and concomitant medications, n (%)** |  |  |  |  |
| Lisinopril | 6,604 (24.9) | 722 (15.0) | 5,882 (27.1) | <0.001 |
| Hydrochlorothiazide | 3,972 (15.0) | 454 (9.4) | 3,518 (16.2) | <0.001 |
| Atorvastatin | 4,132 (15.6) | 395 (8.2) | 3,737 (17.2) | <0.001 |
| Aspirin | 6,104 (23.0) | 589 (12.2) | 5,515 (25.4) | <0.001 |
| Metformin | 4,503 (17.0) | 437 (9.1) | 4,066 (18.7) | <0.001 |
| **Vital signs and care metrics** |  |  |  |  |
| Systolic BP, mean ± SD, mmHg (n) | 140.0 ± 20.6 (20,671) | 141.4 ± 21.4 (4,011) | 139.7 ± 20.4 (16,660) | <0.001 |
| Diastolic BP, mean ± SD, mmHg (n) | 84.7 ± 13.0 (20,671) | 85.6 ± 13.2 (4,011) | 84.5 ± 13.0 (16,660) | <0.001 |
| Treatment rate, n (%; 95% CI) | 9,280 (44.9; 44.2–45.6) | 1,703 (42.5; 40.9–44.0) | 7,577 (45.5; 44.7–46.2) | <0.001 |
| BP control rate, n (%; 95% CI) | 2,851 (10.7; 10.4–11.1) | 413 (8.6; 7.8–9.4) | 2,438 (11.2; 10.8–11.6) | 0.001 |

*BMI, body mass index; BP, blood pressure; CI, confidence interval; SD, standard deviation. p-values compare patients with no follow-up versus patients with $\geq 1$ follow-up visit using chi-square or Fisher's exact tests for categorical variables and Welch's t-test or Wilcoxon rank-sum test for continuous variables, as appropriate. Treatment rate is defined as having an active prescription for at least one antihypertensive medication. BP control is defined as systolic BP <140 mmHg and diastolic BP <90 mmHg.*

At baseline, mean systolic BP was 140.0 ± 20.6 mmHg and mean diastolic BP was 84.7 ± 13.0 mmHg (n = 20,671 with at least one valid baseline measurement). Only 44.9% (95% CI 44.2%–45.6%) of patients were on at least one antihypertensive agent at baseline, and only 10.7% (95% CI 10.4%–11.1%) had BP at the <140/90 mmHg target—values consistent with national surveillance estimates and underscoring the substantial unmet need within these primary-care populations.

### Patterns of follow-up and visit frequency

A striking 18.1% (n = 4,811) of patients had no follow-up visit in the EHR after their index hypertension encounter, despite having a documented diagnosis and an indication for ongoing management (**Figure 1**). Among the 21,729 patients (81.9%) with ≥1 follow-up encounter, the median total visit count was 4 (interquartile range [IQR] 2–9) and the distribution was right-skewed, with a long tail of patients having ≥30 visits during the observation window (**Figure 1**). The median time between consecutive visits decreased as patients accumulated more encounters (Pearson r = –0.217, p < 0.001), consistent with a smaller subset of more clinically complex patients receiving denser follow-up over time (**Figure 2c**).

**Figure 1.**
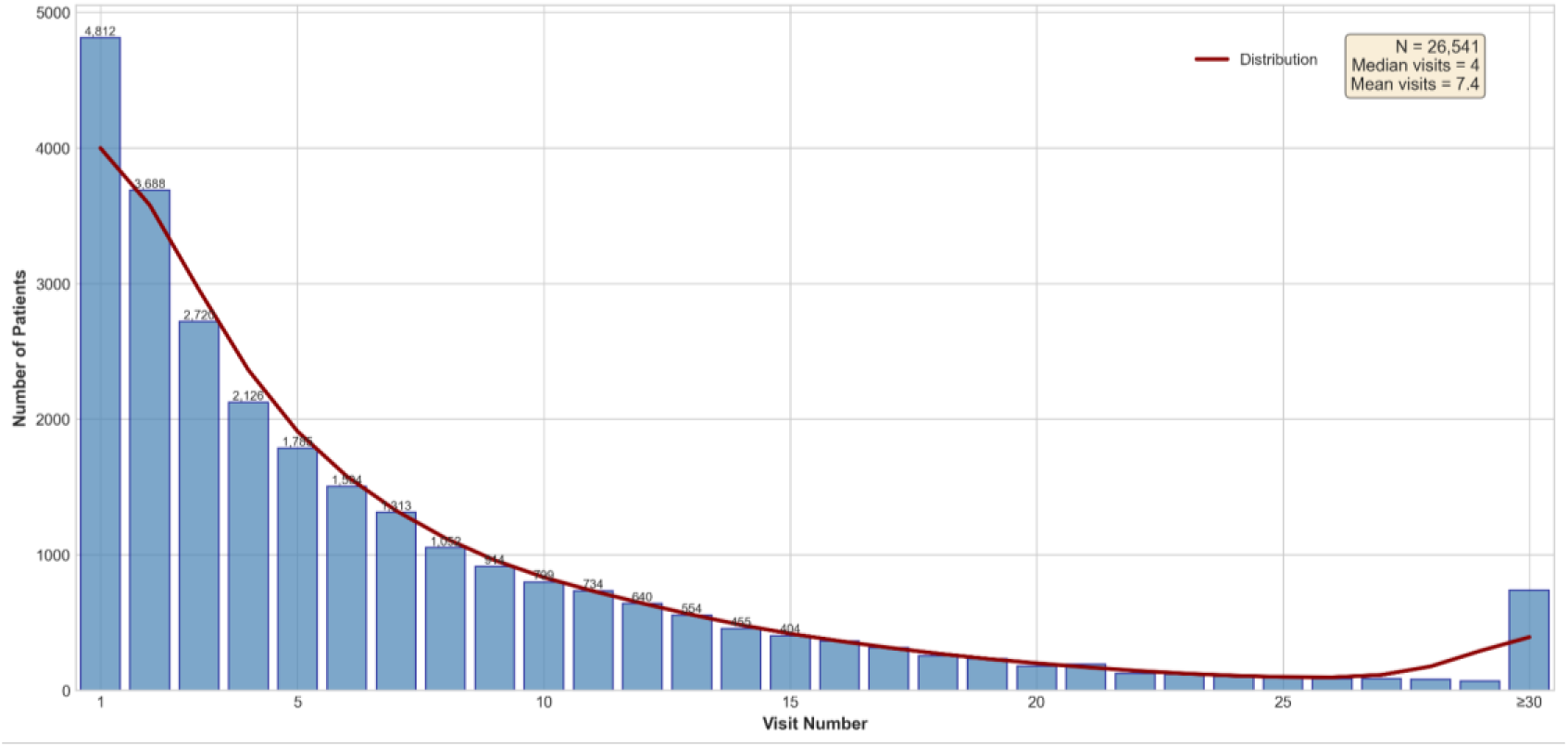
Distribution of total primary-care visits per patient in the EvidenceNOW hypertension cohort. *Histogram of total visit count among 26,541 adults with hypertension. The distribution is right-skewed, with a large concentration of patients having 1–2 visits (median, 4 visits; mean, 7.4) and a long tail of patients with ≥30 visits*.

**Figure 2.**
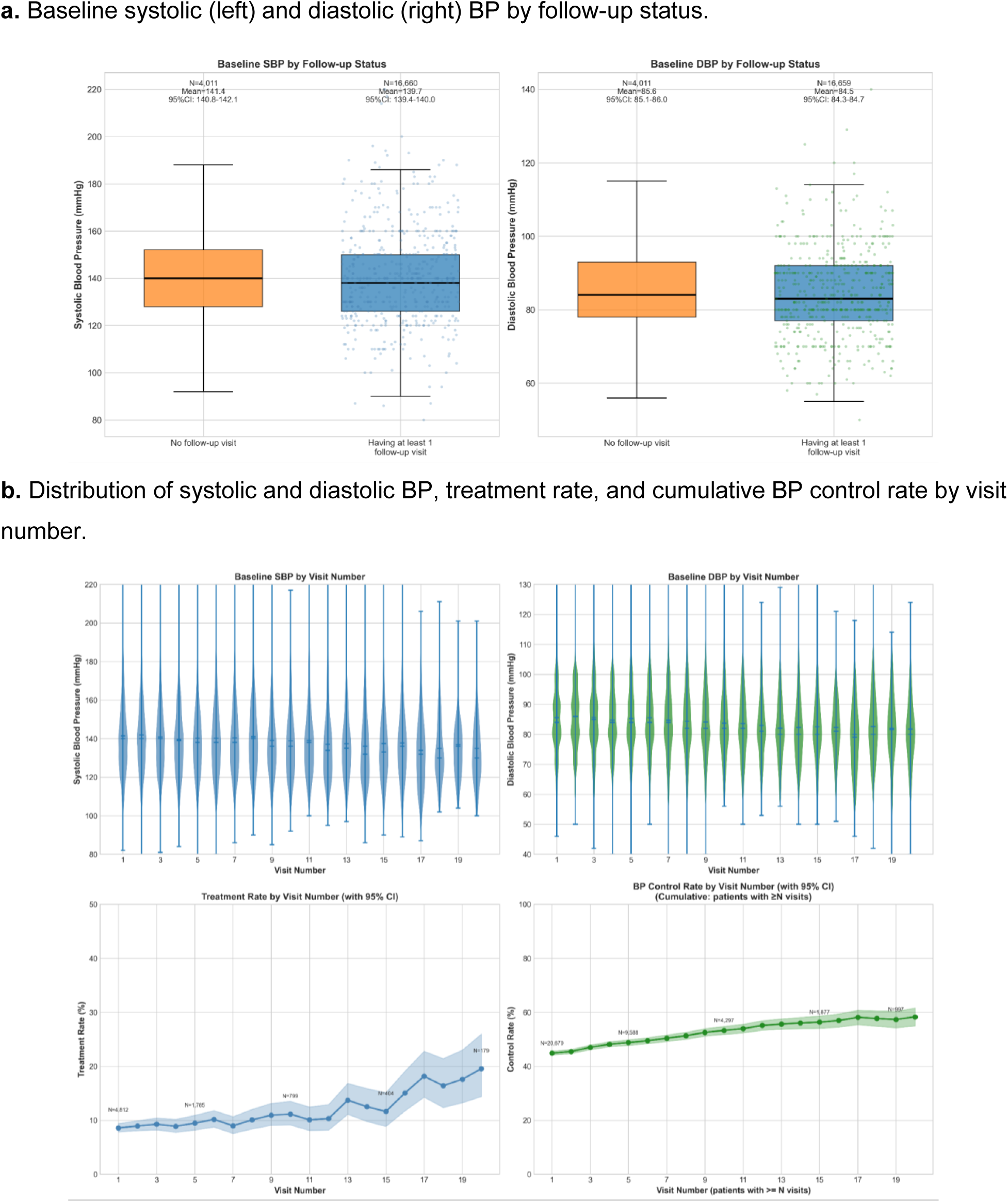
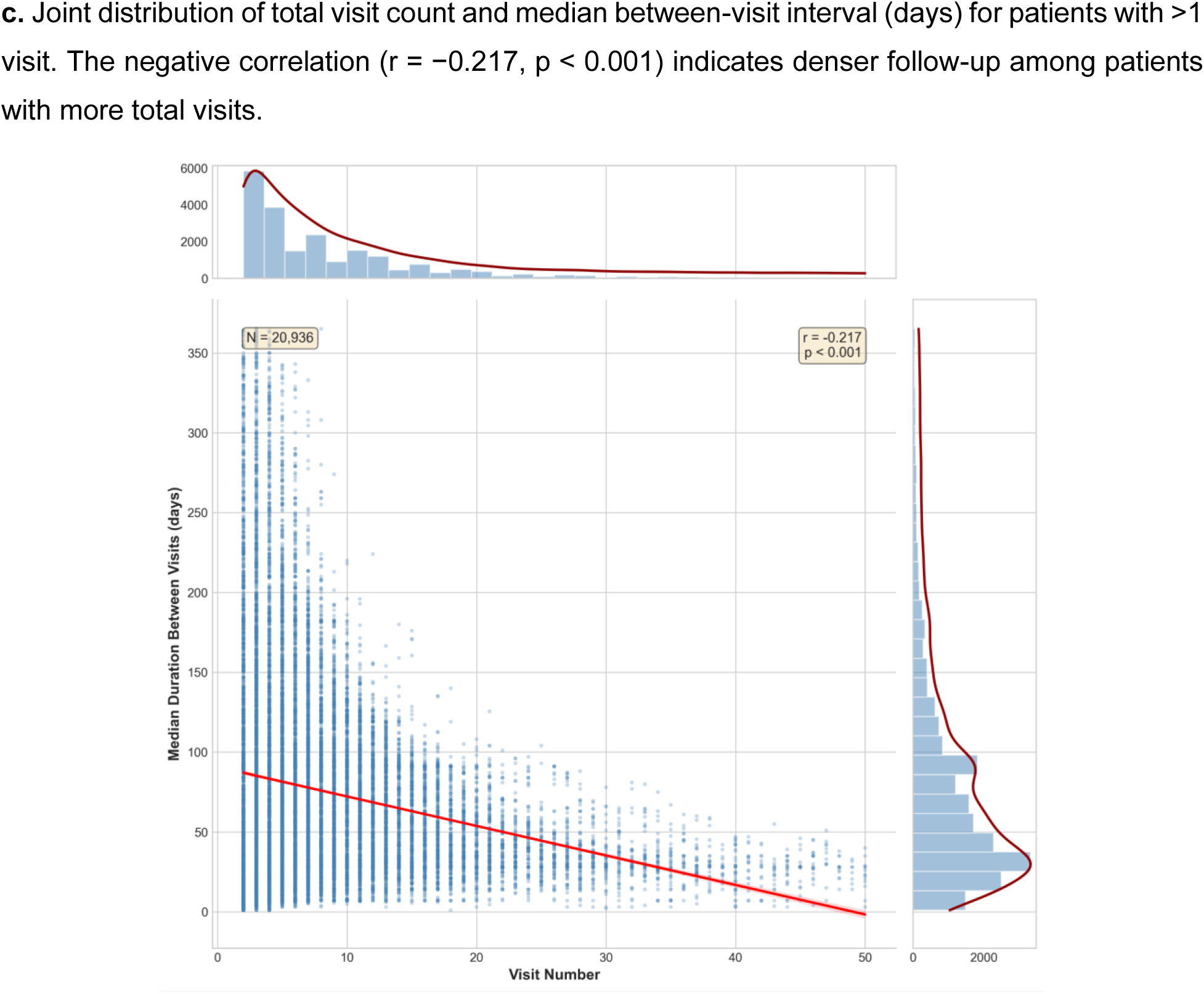
Baseline blood pressure by follow-up status, longitudinal blood pressure trajectories, and the relationship between visit number and time between visits. *Patients without follow-up had higher mean baseline systolic and diastolic BP than those with ≥1 follow-up visit (panel a; both p < 0.001). Among retained patients, mean systolic and diastolic BP improved progressively across visits (panel b, top), accompanied by progressive increases in treatment rate and cumulative BP control rate (panel b, bottom)*.

### Baseline differences between patients with and without follow-up

Patients who never returned for follow-up differed from those who did across several clinically important dimensions (**Table 1**; **Figure 2a**). Patients without follow-up had marginally higher mean age (54.7 vs 54.2 years, p = 0.03) and higher baseline BP—mean SBP 141.4 vs 139.7 mmHg and mean DBP 85.6 vs 84.5 mmHg (both p < 0.001), indicating that those at greatest cardiovascular risk were paradoxically least likely to return. Conversely, patients with follow-up were more likely to be female (56.5% vs 54.9%, p = 0.04), Hispanic/Latino (36.8% vs 30.2%, p < 0.001), or Asian (3.0% vs 1.3%, p < 0.001), and were substantially more likely to have comorbid diabetes (50.3% vs 32.6%), heart failure (4.1% vs 3.3%), chronic kidney disease (4.0% vs 2.4%) or prior stroke (5.4% vs 2.4%) (all p < 0.05). They were also more likely to be on at least one antihypertensive (45.5% vs 42.5%) and to have BP controlled at baseline (11.2% vs 8.6%; p = 0.001).

### Longitudinal retention in care

Among the full cohort of 26,541 patients, the proportion remaining engaged in primary care—operationalized as having at least one additional encounter beyond each index time point—declined steeply over time (**Figure 3** and **Table 2**). Retention was 59.9% at 6 months, 48.2% at 12 months, 39.2% at 18 months, 31.6% at 24 months, 23.8% at 30 months, and 16.3% at 36 months. The shape of the retention curve was monotonically declining without a clear inflection: meaningful attrition was observed at every follow-up interval, suggesting that no single time point dominates loss-to-follow-up risk and that interventions will likely need to operate continuously rather than focusing on a single critical window.

**Figure 3.**
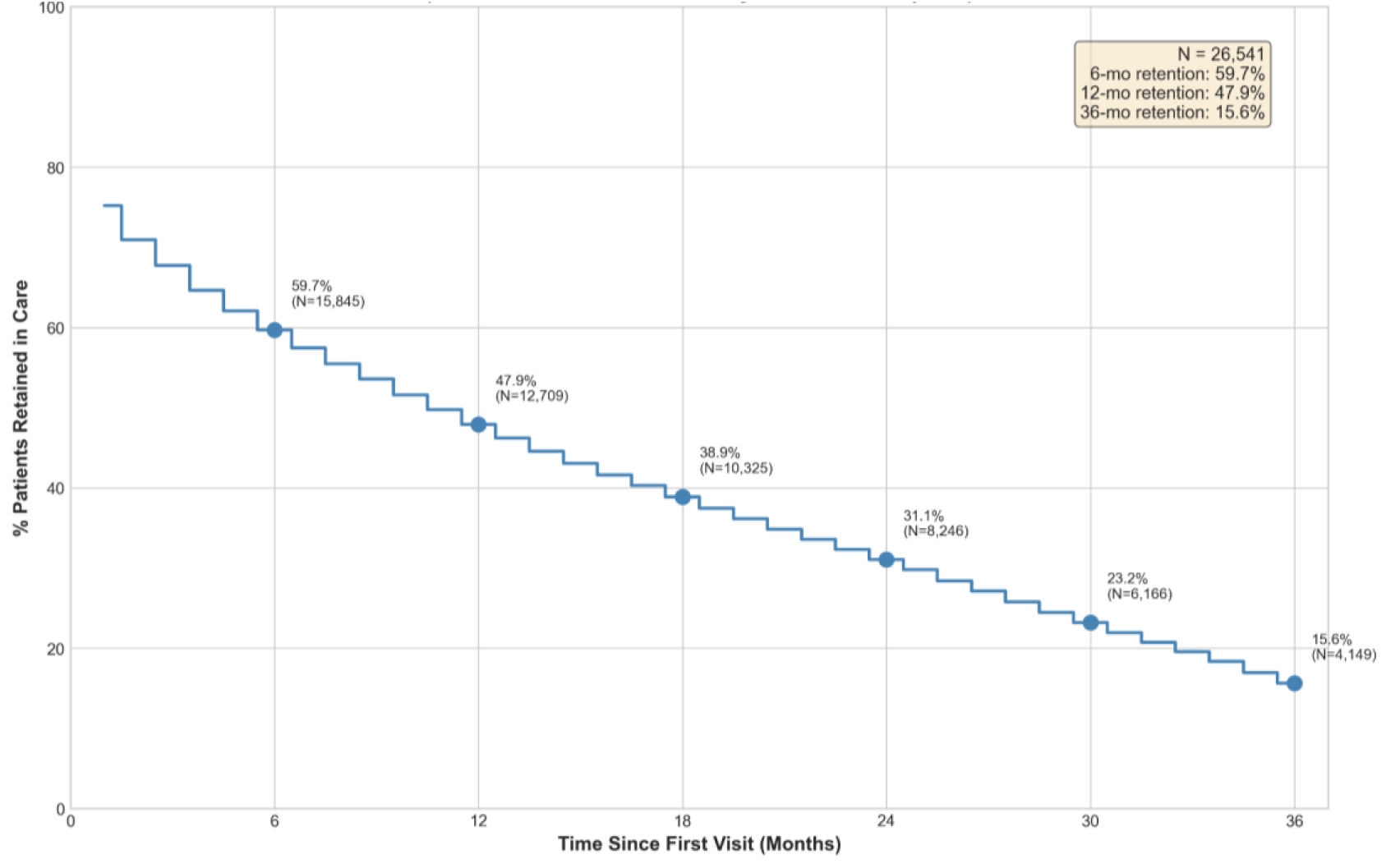
Cumulative retention in primary care over 36 months among patients with hypertension. *Step-function curve showing the proportion of the full cohort (N = 26,541) with at least one primary-care visit beyond each time point. Retention declines monotonically from 59.7% at 6 months to 15.6% at 36 months, with no clear inflection in the curve*.

**Table 2.** Retention in care, care-engagement metrics, and longitudinal blood pressure control among patients with ≥1 follow-up visit.

| Metric | Overall | By age group |  |  |
| --- | --- | --- | --- | --- |
| | | 18–39 y | 40–64 y | $\geq 65$ y |
| **Cumulative retention in care, n (%)\*** |  |  |  |  |
| 6 months | 15,908 (59.9) | 506 (67.7) | 2,109 (70.8) | 1,256 (62.5) |
| 12 months | 12,793 (48.2) | 443 (59.3) | 1,858 (62.4) | 1,016 (50.6) |
| 18 months | 10,411 (39.2) | 397 (53.1) | 1,662 (55.8) | 883 (44.0) |
| 24 months | 8,383 (31.6) | 350 (46.9) | 1,470 (49.4) | 747 (37.2) |
| 30 months | 6,310 (23.8) | 299 (40.0) | 1,284 (43.1) | 641 (31.9) |
| 36 months | 4,323 (16.3) | 243 (32.5) | 1,089 (36.6) | 564 (28.1) |
| **Care engagement metrics, median (IQR)** |  |  |  |  |
| Total visits, n | 4 (2–9) | 6 (2–12) | 7 (3–16) | 7 (2–16) |
| Days to first $\geq 6$-mo gap | 56 (0–204) | 35 (0–156) | 44 (0–182) | 63 (0–236) |
| Days to first $\geq 12$-mo gap | 56 (0–273) | 113 (0–362) | 56 (0–290) | 30 (0–292) |
| Days to first $\geq 18$-mo gap | 33 (0–230) | 115 (0–389) | 37 (0–280) | 14 (0–228) |
| Days from index to last visit | 329 (31–870) | 617 (56–1,240) | 690 (110–1,414) | 372 (51–1,202) |
| Median between-visit interval, days | 58 (29–103) | 49 (23–105) | 46 (26–91) | 34 (25–74) |
| **BP control rate among retained patients, n (%)** |  |  |  |  |
| At 12 months | 1,223 (63.7) | 140 (63.6) | 622 (62.3) | 424 (64.3) |
| At 24 months | 993 (65.8) | 113 (62.4) | 524 (65.2) | 327 (67.1) |
| At 36 months | 775 (64.0) | 80 (62.5) | 439 (64.6) | 237 (63.4) |

BP, blood pressure; IQR, interquartile range. \*Overall retention is calculated relative to the full cohort ( $N = 26,541$ ). Age-stratified retention is calculated among patients within each age stratum who had sufficient potential observation time at the corresponding landmark and therefore should not be summed across strata to obtain the overall figure. BP control rates among retained patients are calculated as the proportion of patients with valid BP measurements at the corresponding landmark whose systolic BP was $<140$ mmHg and diastolic BP was $<90$ mmHg.

Retention varied modestly by age. Among patients with sufficient potential observation time, those aged 40–64 years showed the highest retention at 6 months (70.8%) and 36 months (36.6%), followed by patients aged ≥65 years (62.5% and 28.1%, respectively). The youngest group, aged 18–39 years, had the lowest sustained retention (67.7% at 6 months declining to 32.5% at 36 months). The age gap was relatively small in the early window but widened with extended follow-up, identifying younger adults as a particularly vulnerable subgroup for late attrition.

### Longitudinal BP control among retained patients

Among patients who remained engaged in care, BP control improved markedly relative to baseline. The control rate rose from 11.2% at baseline (among those with ≥1 follow-up) to 63.7% (n = 1,223/1,919) at 12 months, 65.8% (n = 993/1,509) at 24 months, and 64.0% (n = 775/1,211) at 36 months (**Table 2**). Control rates were similar across age strata, with values clustering between 62% and 67% at each landmark. The improvement was paralleled by progressively better mean SBP and DBP at successive visits and by a modest increase in the proportion of patients on at least one antihypertensive agent across visit number (**Figure 2b**)—although treatment rates remained below 30% even at the twentieth encounter, indicating substantial residual room for therapeutic intensification.

Considered together with the retention data, these findings suggest that the dominant lever for population-level BP control in this primary-care setting is not the magnitude of improvement among engaged patients, which is already substantial but rather the proportion of diagnosed patients who remain engaged long enough to achieve it. If the 36-month control rate observed among retained patients could be extended to the full cohort by eliminating loss to follow-up, the absolute population control rate would rise from 10.7% at baseline to a level approaching national targets.

### Predictors of loss to follow-up

We fit a multivariable Cox proportional-hazards model on all 26,540 patients (25,914 events) to identify independent predictors of loss to follow-up after adjusting for demographics, comorbidities, baseline BP control and treatment status, and BMI category (**Figure 4**). Older age (aHR 0.93 per 1-SD increase; 95% CI 0.92–0.94) and female sex (aHR 0.96; 0.93–0.98) were each modestly protective. The strongest demographic signals were observed for race and ethnicity: relative to White patients, Black/African American patients had a 44% higher hazard of loss to follow-up (aHR 1.44; 1.33–1.56), Hispanic/Latino patients a 43% higher hazard (aHR 1.43; 1.35–1.52), and patients of Other/Unknown race a 50% higher hazard (aHR 1.50; 1.41–1.59); the estimate for Asian patients was nonsignificant (aHR 1.08; 0.98–1.18, p = 0.12).

**Figure 4.**
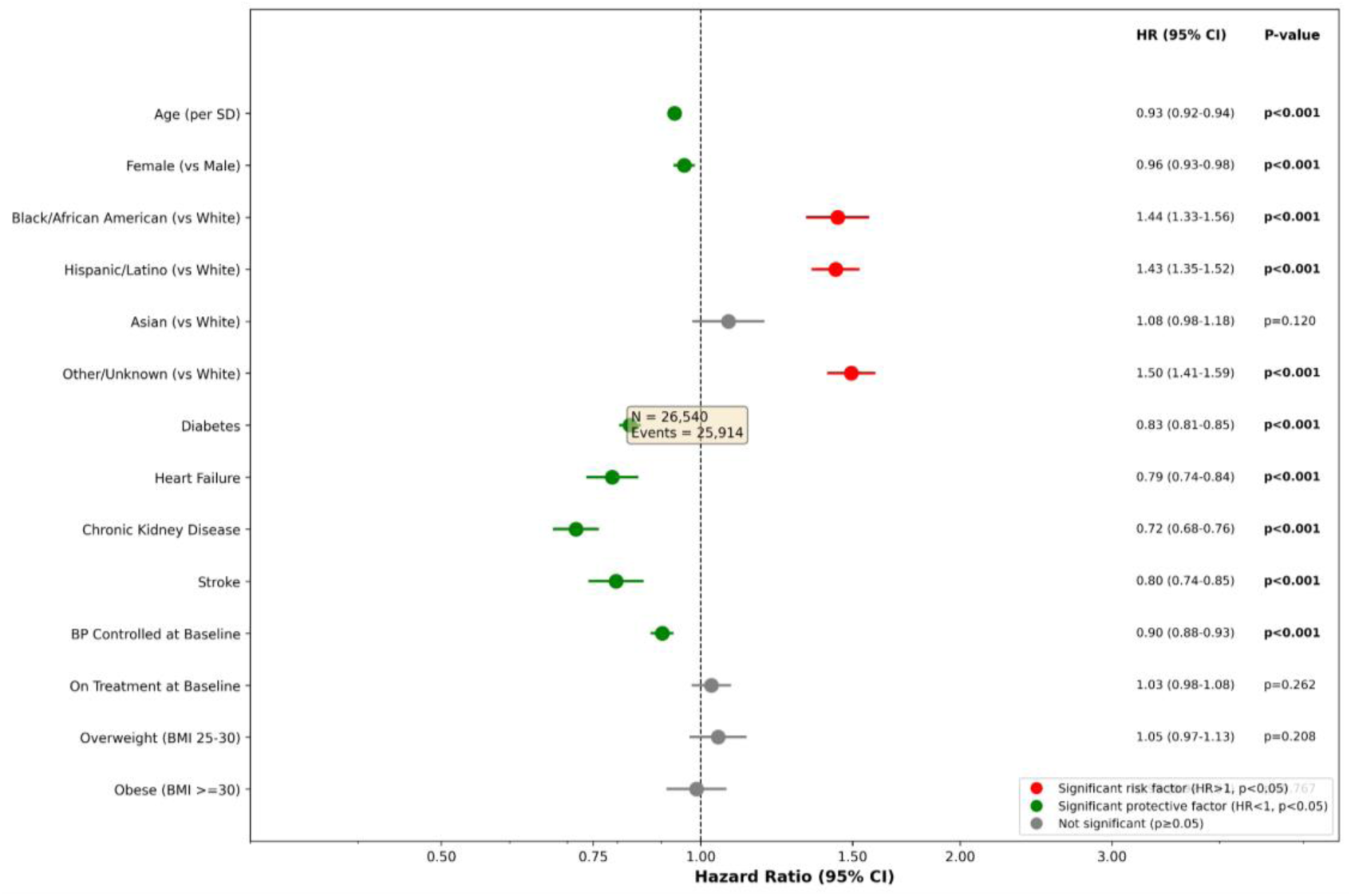
Multivariable Cox proportional-hazards model for loss to follow-up. *Forest plot of adjusted hazard ratios (aHR) and 95% confidence intervals from a multivariable Cox proportional-hazards model in 26,540 patients with 25,914 events. Green points indicate significant protective associations (aHR < 1, p < 0.05); red points indicate significant risk factors (aHR > 1, p < 0.05); grey points indicate non-significant associations. Black/African American, Hispanic/Latino and Other/Unknown race/ethnicity were each independently associated with substantially higher hazards of loss to follow-up after adjustment*.

Comorbidities were uniformly associated with greater retention: diabetes (aHR 0.83; 0.81–0.85), heart failure (aHR 0.79; 0.74–0.84), chronic kidney disease (aHR 0.72; 0.68–0.76) and prior stroke (aHR 0.80; 0.74–0.85) all conferred substantially lower hazards of disengagement, presumably because they require frequent surveillance and reinforce care contact. BP control at baseline was also independently protective (aHR 0.90; 0.88–0.93), whereas baseline antihypertensive treatment did not independently predict retention (aHR 1.03; 0.98–1.08, p = 0.26). Overweight (BMI 25–30) and obese (BMI ≥30) categories were not significantly associated with loss to follow-up after adjustment.

## DISCUSSION

In this multi-practice cohort of 26,541 patients with hypertension, we observed two simultaneous and complementary findings that together explain a large fraction of the persistent national gap in BP control.[2] First, the trajectory of patients through real-world primary care is dominated by attrition: nearly one in five patients never returned after diagnosis, and only one in six remained engaged at three years. Second, among patients who did remain engaged, BP control rose from 11% at baseline to roughly 64% by 12 months and stayed at that level through 36 months. These two patterns, taken together, identify retention, rather than the depth of management once a patient is engaged, as the principal limiting factor for population-level hypertension control in this setting.[11]

The retention curves we report are sobering. They are also, in their shape, inconsistent with a model in which loss to follow-up is dominated by a single early dropout window followed by a stable cohort of engaged patients. Instead, attrition was monotonic and substantial at every observation interval. Practical implications follow directly: outreach interventions, including text-message reminders, panel-management nurse calls, community healthcare worker programs,[15] may be unlikely to succeed if they are delivered only at a single landmark (e.g., 30 days post-diagnosis) and instead need to operate continuously.[7] The plateau of BP control at ≈64% among retained patients, despite ongoing engagement, also suggests that retention alone is insufficient: a meaningful subset of consistently engaged patients are not having their regimen intensified to target, in keeping with the long-described phenomenon of clinical inertia[16] and with reports of difficult-to-control or apparent treatment-resistant hypertension in primary-care populations.[17]

The most striking signals in our analysis were the racial and ethnic disparities in retention. Compared with White patients, Black/African American, Hispanic/Latino, and Other/Unknown-race patients all had 43–50% higher adjusted hazards of loss to follow-up.[18] These differences persisted after adjustment for age, sex, comorbidities, BMI, and baseline BP control. Although our data cannot adjudicate underlying mechanisms, these gaps almost certainly reflect a combination of structural factors including language access, immigration related fears about healthcare contact,[12] transportation, work schedules,[19] insurance discontinuity, and trust in the health system rather than individual patient choice.[20] They demand that retention-focused interventions be designed and evaluated explicitly through an equity lens,[21] with success measured by reductions in disparities and not solely by mean improvement.[10]

The protective association between comorbidities and retention is clinically intuitive but has important consequences. Patients with diabetes, heart failure, chronic kidney disease, or prior stroke are anchored in care by other diseases that demand contact; their hypertension benefits from spillover engagement. The corollary is that patients with isolated, asymptomatic hypertension, who arguably represent the largest opportunity for primary prevention, lack this anchoring effect and are most vulnerable to disengagement.[22] They are also, in our data, the patients most likely to begin care with elevated BP and least likely to be on therapy. Targeting this group with proactive outreach, medication starter sets, and home BP monitoring may have a disproportionate population-level impact.

### Limitation

This study has several limitations. First, EvidenceNOW data come from primary care practices and do not capture care delivered elsewhere; some patients we classified as lost to follow-up may have transferred to non-participating clinicians or moved out of catchment, which may results in conservative estimates of retention. Second, the high proportion of patients categorized as “Other” race/ethnicity (50.5%) reflects well-known limitations of EHR demographic capture and may obscure heterogeneity within minoritized groups; future work using self-reported, granular categories is needed. Third, our analyses are observational and cannot support causal claims about the effects of any single demographic or clinical factor.[20] Notwithstanding these limitations, our findings carry concrete and timely implications. They reframe the population-level hypertension-control problem as primarily an engagement problem, identify specific demographic groups for whom engagement gaps are largest, and quantify the magnitude of improvement attainable when patients are simply retained. They also point to a tractable evaluation framework: retention at fixed landmarks (6-, 12-, 24-, 36- months) and disparities in retention should be reported alongside, and weighted equally with, BP control as primary endpoints in hypertension quality-improvement programs.[13] Pairing the prescribing improvements made possible by guideline implementation with explicit, equity-focused retention strategies offers the most promising path toward meaningful,[23] durable progress against the leading modifiable cause of cardiovascular death.[3]

## CONCLUSION

This multi-practice cohort study demonstrates that the predominant barrier to population-level hypertension control in U.S. primary care is not inadequate treatment among engaged patients but rather the failure to retain patients in care over time. The steep, unrelenting decline in retention, compounded by pronounced racial and ethnic disparities in disengagement, means that even effective prescribing practices reach only a shrinking fraction of the diagnosed population. Critically, patients who remained engaged achieved and sustained BP control rates exceeding 60%, proving that existing primary-care infrastructure is already capable of delivering meaningful clinical benefit when continuity is maintained. These findings call for a fundamental shift in how hypertension quality improvement is conceived: from a narrow focus on treatment intensification at the point of care to a broader, equity-centered strategy that treats longitudinal patient retention as a measurable, reportable, and improvable outcome.

### Author contributions

JY designed the study. JY and AS led the data extraction and statistical analysis. JY wrote the manuscript. All authors contributed to interpretation of the results, critical revision of the manuscript for important intellectual content, and approved the final version for submission.

## Acknowledgments

We thank the participating EvidenceNOW practices, clinicians and quality-improvement staff for their commitment to improving cardiovascular preventive care in their communities, and the patients whose deidentified data made this work possible.

## Data availability

Datasets are available from the corresponding author on reasonable request and completion of appropriate data sharing agreements. The relevant code and analyses are available at https://github.com/sjcalan/Hypertension.

## REFERENCES

1. Mills, K.T., A. Stefanescu, and J. He, The global epidemiology of hypertension. Nature Reviews Nephrology, 2020. 16(4): p. 223–237.

2. Hardy, S.T., et al., Trends in blood pressure control among US adults with hypertension, 2013–2014 to 2021–2023. American Journal of Hypertension, 2025. 38(2): p. 120–128.

3. Frieden, T.R. and M.G. Jaffe, Saving 100 million lives by improving global treatment of hypertension and reducing cardiovascular disease risk factors. The Journal of Clinical Hypertension, 2018. 20(2): p. 208.

4. Ye, J., S. Bronstein, and H. Chen, WHO HEARTS Programs in High-Income Countries: Implementation, Outcomes, and Implications for Global Hypertension Control. American Journal of Preventive Cardiology, 2026: p. 101579.

5. Reboussin, D.M., et al., Systematic review for the 2017 ACC/AHA/AAPA/ABC/ACPM/AGS/APhA/ASH/ASPC/NMA/PCNA guideline for the prevention, detection, evaluation, and management of high blood pressure in adults: a report of the American College of Cardiology/American Heart Association Task Force on Clinical Practice Guidelines. Journal of the American College of Cardiology, 2018. 71(19): p. 2176–2198.

6. Ye, J., et al., Optimizing Longitudinal Retention in Care Among Patients With Hypertension in Primary Healthcare Settings: Findings From the Hypertension Treatment in Nigeria Program. Circulation, 2022. 146(Suppl_1): p. A13217–A13217.

7. Kronish, I.M., et al., Meta-analysis: impact of drug class on adherence to antihypertensives. Circulation, 2011. 123(15): p. 1611–1621.

8. Ye, J., et al., Characteristics and Patterns of Retention in Hypertension Care in Primary Care Settings From the Hypertension Treatment in Nigeria Program. JAMA Network Open, 2022. 5(9): p. e2230025–e2230025.

9. Chen, H. and J. Ye, Demographic Patterns, Social Determinants of Health, and Climate Change Perceptions Among Patients with Cancer and Hypertension. medRxiv, 2026: p. 2026.01. 21.26344565.

10. Ye, J., et al., Interventions and contextual factors to improve retention in care for patients with hypertension in primary care: Hermeneutic systematic review. Preventive Medicine, 2024: p. 107880.

11. Jaffe, M.G., et al., Improved blood pressure control associated with a large-scale hypertension program. Jama, 2013. 310(7).

12. Chen, H., et al., Telehealth Utilization and Patient Experiences: The Role of Social Determinants of Health Among Individuals with Hypertension and Diabetes. medRxiv, 2024: p. 2024.08. 01.24311392.

13. Meyers, D., et al., EvidenceNOW: balancing primary care implementation and implementation research. The Annals of Family Medicine, 2018. 16(Suppl 1): p. S5–S11.

14. Abdalla, M., et al., Implementation strategies to improve blood pressure control in the United States: a scientific statement from the American Heart Association and American Medical Association. Hypertension, 2023. 80(10): p. e143–e157.

15. Ye, J., et al., Community-Based Participatory Research and System Dynamics Modeling for Improving Retention in Hypertension Care. JAMA Network Open, 2024. 7(8): p. e2430213–e2430213.

16. Phillips, L.S., et al., Clinical inertia. Annals of internal medicine, 2001. 135(9): p. 825–834.

17. Daugherty, S.L., et al., Incidence and prognosis of resistant hypertension in hypertensive patients. Circulation, 2012. 125(13): p. 1635–1642.

18. Abrahamowicz, A.A., et al., Racial and ethnic disparities in hypertension: barriers and opportunities to improve blood pressure control. Current Cardiology Reports, 2023. 25(1): p. 17–27.

19. Chen, H. and J. Ye, Digital Health Technology Burden and Frustration Among Patients with Multimorbidity. medRxiv, 2025: p. 2025.10. 09.25337645.

20. Bress, A.P., et al., Inequities in hypertension control in the United States exposed and exacerbated by COVID-19 and the role of home blood pressure and virtual health care during and after the COVID-19 pandemic. 2021.

21. Hashish, M.A., S. Bronstein, and J. Ye, Returning value to communities from the All of Us Research Program through the lens of social determinants of health and ethical, legal, and social implications. International Journal for Equity in Health, 2026.

22. Ye, J., et al., Implementation outcomes from the Hypertension Treatment in Nigeria program: results from a type 2 hybrid interrupted time series trial. Implementation Science, 2025.

23. Carey, R.M., et al., Reprint of: prevention and control of hypertension: JACC Health Promotion Series. Journal of the American College of Cardiology, 2018. 72(23 Part B): p. 2996–3011.

